# Trends and patterns of dual use of combustible tobacco and e-cigarettes among adults in England: a population study, 2016-2024

**DOI:** 10.1101/2024.07.17.24310557

**Authors:** Sarah E. Jackson, Sharon Cox, Lion Shahab, Jamie Brown

**Author notes:** Corresponding author: Dr Sarah Jackson, Department of Behavioural Science and Health, University College London, 1-19 Torrington Place, London WC1E 7HB, UK. **Declaration of interests:** JB has received unrestricted research funding from Pfizer and J&J, who manufacture smoking cessation medications. LS has received honoraria for talks, unrestricted research grants and travel expenses to attend meetings and workshops from manufactures of smoking cessation medications (Pfizer; J&J), and has acted as paid reviewer for grant awarding bodies and as a paid consultant for health care companies. All authors declare no financial links with tobacco companies, e-cigarette manufacturers, or their representatives. **Pre-registration:** https://osf.io/pqadf/.

## Abstract

**Background/Aims:** E-cigarettes are frequently used by people who smoke. This study examined how the prevalence and patterns of smoking and vaping (‘dual use’) in England have changed as the vaping market has rapidly evolved.

**Design:** Representative monthly cross-sectional survey, July 2016 to April 2024.

**Setting:** England.

**Participants:** 128,588 adults (≥18y).

**Measurements:** Logistic regression estimated associations between survey wave and dual use. Descriptive statistics were used to analyse patterns of smoking and vaping, overall and by sociodemographic, smoking, and vaping characteristics and harm perceptions of e-cigarettes vs. cigarettes.

**Results:** Across the period, the overall prevalence of dual use increased non-linearly from 3.5% to 5.3% of adults (prevalence ratio [PR]=1.49 [1.25-1.76]). Among adults who smoked, the proportion who also vaped was relatively stable up to mid-2021, at an average of 18.6% between July-2016 and May-2021, then increased rapidly to 34.2% by April 2024 (PR=1.76 [1.48-2.09]). This increase was greatest at younger ages (e.g., from 19.6% to 59.4% among 18-24-year-olds; PR=3.04 [2.28-4.23]). The most common pattern of dual use across the period was daily cigarette smoking with daily vaping (49.0% [47.3-50.8%]). Over time, the proportion of dual users reporting daily cigarette smoking with non-daily vaping decreased (from 35.2% to 15.0%; PR=0.43 [0.29-0.63]), offset primarily by an increase in the proportion reporting non-daily cigarette smoking with daily vaping (from 7.6% to 21.5%; PR=2.84 [1.71-4.72]). Daily cigarette smoking with daily vaping was more common (and non-daily cigarette smoking with daily vaping less common) among dual users who were older, less advantaged, mainly smoked hand-rolled cigarettes, had stronger urges to smoke, and had been vaping for ≤6 months. Daily vaping was more common among dual users who thought e-cigarettes were less/equally harmful as cigarettes, or were unsure.

**Conclusions:** The proportion of smokers in England who vape has increased rapidly since 2021, which was when disposable e-cigarettes started to become popular. Since 2016, patterns of dual use have shifted away from more frequent smoking towards more frequent vaping. This may be the result of increasing prevalence of dual use among younger adults, who are more likely than older dual users to smoke non-daily and vape daily.

## Introduction

E-cigarettes are frequently used by people who smoke combustible tobacco. For some, dual use (use of an e-cigarette alongside smoking) may be a temporary state as they transition away from smoking. Many smokers say they use e-cigarettes (vape) to reduce the amount they smoke or to aid smoking cessation^1^ and substantial evidence shows e-cigarettes are effective for helping people to stop smoking.^2,3^ For others, dual use may be motivated by other reasons, for example, because they enjoy vaping or because it offers a means to use nicotine in situations where smoking is not permitted.^1,4^ Given vaping is much less harmful than smoking,^5^ the health implications of dual use will depend on the frequency with which a person engages in each behaviour. A person who vapes every day and smokes occasionally is likely to be exposed to lower levels of potential toxicants than someone who smokes and vapes daily.^6^ Understanding trends in the prevalence and patterns of dual use – and how they differ according to relevant sociodemographic, smoking, and vaping characteristics – can inform public health messaging around smoking and vaping.

In England, representative household surveys suggest the proportion of adult smokers who vape has increased over recent years. In 2014-16, around one in four (23.7%) adults who smoked either daily or non-daily also vaped.^7^ By 2023, this number had risen to around one in three (31.5%).^8^ It is not clear whether there has been a corresponding increase in the overall prevalence of dual use among adults (given the proportion of adults who smoke has fallen over this period^9^), whether the increase in vaping among smokers has been linear or predominantly occurred in the last few years (vaping prevalence in the adult population was relatively stable between 2014 and 2020 but has increased rapidly since 2021^10,11^), or the extent to which it has been observed to a similar extent across smokers with different sociodemographic characteristics.

In terms of patterns of dual use, data from the International Tobacco Control (ITC) Survey indicated that around half (52.4%) of dual users in England in 2016 predominantly smoked (i.e., smoked daily and vaped non-daily); just one in ten (9.7%) predominantly vaped (i.e., vaped daily and smoked non-daily).^12^ It is not clear whether this has changed over time in the context of declining rates of smoking^9^ and recent increases in vaping.^10,11^ There has also been a recent increase in non-cigarette tobacco smoking (e.g., cigars, cigarillos, or shisha) in England since 2020,^13^ and there is little data on the intersection between non-cigarette smoking and vaping. Patterns of dual use may differ according to the characteristics of the user,^12^ such as their sociodemographic background, the types of combustible tobacco products they use, how dependent they are on smoking, their reasons for vaping, how long they have been vaping for, and how they perceive the relative harms of the two products. For example, older adults who dual use may be more likely to smoke daily than younger dual users as their smoking habits and routines may be more entrenched, whereas younger adults may see smoking and vaping as more interchangeable.^14^ People who have been vaping for longer may transition away from more regular smoking towards more regular vaping, and those who perceive e-cigarettes as less harmful than smoking may be more likely to vape more regularly than they smoke compared with those who perceive the harms to be similar or who believe vaping is more harmful.

This study aimed to understand more about the prevalence and patterns of dual use of combustible tobacco and e-cigarettes among adults in England. Using data from a nationally-representative survey, we estimated time trends in the overall prevalence of dual use among (i) adults and (ii) smokers in England between 2016 and 2024, overall and within sociodemographic subgroups. We also described patterns of smoking (daily cigarette vs. non-daily cigarette vs. non-cigarette tobacco) and vaping (daily vs. non-daily) among dual users over this period, overall and by sociodemographic, smoking, and vaping characteristics and harm perceptions of e-cigarettes vs. cigarettes.

## Methods

### Pre-registration

The study protocol, research questions, and analysis plan were pre-registered on Open Science Framework (https://osf.io/pqadf/).

### Design

We analysed data from the Smoking Toolkit Study, an ongoing representative repeat cross-sectional survey of adults in England.^15^ Each month, a new sample of approximately 1,700 adults (≥16y) is selected via a hybrid of random probability and simple quota sampling. Data were collected face-to-face up to the start of the Covid-19 pandemic and have been collected via telephone interviews since April 2020; the two modalities show good comparability on key sociodemographic and nicotine use indices.^16^

The present analyses focused on data collected between July 2016 (the first wave to collect information on vaping frequency) and April 2024 (the most recent data on vaping frequency at the time of analysis). Vaping characteristics were not assessed in certain waves during this period (May, June, August, September, November, and December 2022; February, March, May, August, September, November, and December 2023; and February, and March 2024), so we excluded participants surveyed in these waves. Data were not collected from 16- and 17-year olds between April 2020 and December 2021, so we restricted the sample to those aged 18 and older for consistency across the period.

### Measures

#### Smoking status and frequency

Smoking status was assessed by asking participants which of the following best applied to them:

(a) I smoke cigarettes (including hand-rolled) every day; (b) I smoke cigarettes (including hand-rolled), but not every day; (c) I do not smoke cigarettes at all, but I do smoke tobacco of some kind (e.g., pipe, cigar or shisha); (d) I have stopped smoking completely in the last year; (e) I stopped smoking completely more than a year ago; or (f) I have never been a smoker (i.e., smoked for a year or more). Those who responded *a* or *b* were considered current cigarette smokers. Those who responded *c* were considered exclusive non-cigarette tobacco smokers. Cigarette smoking frequency was categorised according to responses to the same question as daily (response *a*) and non-daily (response *b*). Frequency of non-cigarette smoking was not captured by the survey.

#### Vaping status and frequency

Vaping status was assessed among current smokers within three questions that asked about use of a range of nicotine products: ‘Do you regularly use any of the following in situations when you are not allowed to smoke?’; ‘Which, if any, of the following are you currently using to help you cut down the amount you smoke?’; and ‘Can I check, are you using any of the following either to help you stop smoking, to help you cut down or for any other reason at all?’. Those who reported using an e-cigarette in response to any of these questions were considered current vapers. Other response options included different types of nicotine replacement therapy, heated tobacco products, and nicotine pouches.

To assess vaping frequency, participants who vaped were asked: ‘How many times per day on average do you use your nicotine replacement product or products?’ Response options were 1, 2, 3-4, 5-7, 8-11, 12+, not every day but at least once a week, not everyday and less than once a week, and don’t know. Those who reported use at least once a day were considered to be vaping daily and those who reported use less than once a day were considered to be vaping non-daily. We treated those who responded that they did not know as missing.

#### Dual use

Those who reported current smoking and current vaping were considered dual users. Within this group, we classified patterns of dual use as:

a. Daily cigarette smoking with daily vaping
b. Daily cigarette smoking with non-daily vaping
c. Non-daily cigarette smoking with daily vaping
d. Non-daily cigarette smoking with non-daily vaping
e. Non-cigarette tobacco smoking with daily vaping
f. Non-cigarette tobacco smoking with non-daily vaping

Because the measure assessing vaping frequency was not specific to vaping, we conducted a sensitivity analysis restricting the sample to those reporting no current use of nicotine replacement therapy, heated tobacco products, or nicotine pouches.

#### Sociodemographic characteristics

Age was categorised as 18-24, 25-34, 35-44, 45-54, 55-64, and ≥65 years. Gender was self-reported as man or woman (in more recent waves, participants have also had the option to describe their gender in another way; those who identified in another way were excluded from analyses by gender due to low numbers). Occupational social grade was categorised as ABC1 (includes managerial, professional, and upper supervisory occupations) and C2DE (includes manual routine, semi-routine, lower supervisory, state pension, and long-term unemployed).

#### Smoking characteristics

Strength of urges to smoke was assessed with self-reported ratings of urges over the past 24 hours, with the response options: not at all, slight, moderate, strong, very strong and extremely strong. We also included within the ‘not at all’ category those who responded ‘not at all’ to the (separate) question: ‘How much of the time have you spent with the urge to smoke?’.^17^

The main type of cigarettes smoked was assessed among current cigarette smokers by asking how many cigarettes they usually smoked (per day or week, as preferred) and how many of these were hand-rolled. We classified the main type of cigarettes smoked as hand-rolled for those reporting at least 50% of their total cigarette consumption was hand-rolled and manufactured for those reporting that less than 50% was hand-rolled. The type of non-cigarette combustible tobacco products used was not captured by the survey.

#### Vaping characteristics

Duration of vaping was assessed among current vapers with the question: ‘How long have you been using this nicotine replacement product or these products for?’ We distinguished between those who had been vaping for ≤6 months, >6 and ≤12 months, and >12 months.

Vaping purpose was defined according to responses to the questions assessing current vaping, as described above. We analysed those reporting vaping for: (i) cutting down the amount smoked, (ii) use in situations where smoking is not permitted, and (iii) other reasons (i.e., those who did not report vaping for either of these reasons). Note that (i) and (ii) were not mutually exclusive. Each of these three reasons for vaping was analysed as a separate binary variable.

Use of e-cigarettes in a past-year quit attempt as assessed by asking: ‘How many serious attempts to stop smoking have you made in the last 12 months? By serious attempt I mean you decided that you would try to make sure you never smoked again. Please include any attempt that you are currently making and please include any successful attempt made within the last year’. Those who reported having made at least one quit attempt were then asked: ‘What did you use to help stop smoking during the most recent serious quit attempt?’ Those who responded ‘electronic cigarette’ were considered to have used an e-cigarette in a past-year quit attempt. Those who either did not report a quit attempt or did not report using an e-cigarette in their most recent quit attempt were considered not to have used an e-cigarette in a past-year quit attempt.

#### Harm perceptions of e-cigarettes vs. cigarettes

Harm perceptions were assessed with the question: ‘Compared to regular cigarettes, do you think electronic cigarettes are more, less, or equally harmful to health?’ Response options were ‘more harmful’, ‘less harmful’, ‘equally harmful’, or ‘don’t know’. We dummy coded these response options as one-versus-else (e.g., less harmful vs. all other responses) for analysis.^18^

### Statistical analysis

Data were analysed using R v.4.2.1. The Smoking Toolkit Study uses raking to weight the sample to match the population in England.^15^ The following analyses used weighted data (we report all sample sizes as unweighted). We excluded participants with missing data on smoking or vaping status. Missing cases on other variables (consistently <5%) were excluded on a per-analysis basis.

#### Trends in the overall prevalence of dual use

We used logistic regression to estimate time trends in the overall prevalence of dual use among (i) adults and (ii) smokers across the study period, with dual use as the outcome and time (survey month) modelled using restricted cubic splines with five knots. This allowed changes over time to be flexible and non-linear.

To explore moderation of trends by age, gender, and occupational social grade, we repeated models including the interaction between the moderator of interest and time – thus allowing for time trends to differ across subgroups. Each of the interactions was tested in a separate model.

We used predicted estimates to plot the prevalence of dual use over the study period, overall among adults and smokers and by sociodemographic characteristics. We also calculated prevalence ratios (PRs) for the change in prevalence from the start to the end of the period, alongside 95% confidence intervals (CIs) calculated using bootstrapping (1,000 replications).

For context, we also plotted unmodelled datapoints showing the proportion of adults who reported dual use versus exclusive smoking, to illustrate how changes in dual use have occurred in the context of overall changes in smoking prevalence (unplanned analysis).

#### Patterns of dual use across the study period

Using data aggregated across survey waves, we used descriptive statistics to report the percentage (and 95% CI) of adult dual users who reported each pattern of dual use, overall and by sociodemographic, smoking, and vaping characteristics, and harm perceptions (as described in the *measures* section). We based our interpretation of differences in patterns of dual use between subgroups on 95% CIs.

To explore changes in patterns of dual use among dual users across the period, we used logistic regression to analyse the association between each dual use pattern (as the outcome; dummy coded) and time (survey month; modelled using restricted cubic splines). We used predicted estimates to plot the prevalence of each dual use pattern over the study period. We also plotted unmodelled datapoints showing patterns of dual use by survey year (coded as 12-month periods from July-June; e.g., 2016/17 = July 2016 to June 2017) among (i) adults, (ii) smokers, and (iii) dual users.

As a sensitivity analysis, we repeated the descriptive analysis of patterns of dual use (overall) restricting the sample of dual users to those not also using nicotine replacement therapy, heated tobacco products, or nicotine pouches. Given the pattern of results was very similar, we did not extend this sensitivity analysis to the subgroup or trend analyses.

## Results

A total of 129,038 (unweighted) adults (≥18y) in England were surveyed in eligible waves. We excluded 450 (0.3%) with missing data on smoking or vaping status. This resulted in a sample of 128,588 adults for analysis (weighted mean [SD] age = 48.1 [18.7] years; 50.8% women; 44.3% social grades C2DE), of whom 20,749 (16.1%) were smokers (weighted mean [SD] age = 42.3 [16.6] years; 46.3% women; 59.6% social grades C2DE).

### Trends in the overall prevalence of dual use

Between July 2016 and April 2024, the overall prevalence of dual use (i.e., the proportion of adults who both smoked and vaped) increased from 3.5% to 5.3% (PR=1.49 [1.25–1.76]; **Table 1**). This was driven by an increase in vaping rather than smoking: smoking prevalence declined non-linearly over this period (**Figure S1**).

**Table 1.** Modelled estimates of the overall prevalence of dual use of smoking and vaping in July 2016 and April 2024.

|  | Adults |  |  |  | Smokers |  |  |  |
| --- | --- | --- | --- | --- | --- | --- | --- | --- |
|  | N<sup>1</sup> | Prevalence, % [95%CI]<sup>2</sup> |  | Prevalence ratio [95%CI]<sup>3</sup> | N<sup>1</sup> | Prevalence, % [95%CI]<sup>2</sup> |  | Prevalence ratio [95%CI]<sup>3</sup> |
|  |  | July 2016 | April 2024 |  |  | July 2016 | April 2024 |  |
| Overall | 128,588 | 3.5 [3.1–4.0] | 5.2 [4.6–5.9] | 1.49 [1.25–1.76] | 20,749 | 19.5 [17.4–21.7] | 34.2 [30.9–37.8] | 1.76 [1.52–2.04] |
| Age (years) |  |  |  |  |  |  |  |  |
| 18-24 | 15,563 | 4.7 [3.5–6.2] | 9.9 [7.7–12.7] | 2.13 [1.49–3.11] | 3,406 | 19.6 [14.8–25.4] | 59.4 [50.0–68.2] | 3.04 [2.28–4.23] |
| 25-34 | 18,692 | 4.4 [3.3–5.8] | 7.8 [6.0–10.0] | 1.78 [1.22–2.57] | 4,361 | 19.5 [14.9–25.1] | 39.4 [31.9–47.5] | 2.02 [1.52–2.88] |
| 35-44 | 18,174 | 4.4 [3.3–5.8] | 5.6 [4.1–7.6] | 1.28 [0.84–1.98] | 3,328 | 23.3 [17.9–29.8] | 33.6 [26.0–42.2] | 1.44 [1.02–2.16] |
| 45-54 | 20,578 | 3.9 [3.0–5.2] | 5.8 [4.2–7.8] | 1.46 [0.97–2.19] | 3,507 | 19.3 [14.9–24.5] | 31.9 [24.2–40.8] | 1.66 [1.17–2.37] |
| 55-65 | 20,861 | 3.0 [2.2–4.1] | 3.2 [2.3–4.5] | 1.08 [0.71–1.61] | 3,028 | 17.7 [13.2–23.3] | 20.0 [14.4–27.2] | 1.13 [0.73–1.67] |
| ≥65 | 34,720 | 1.5 [1.1–2.2] | 1.5 [1.0–2.5] | 1.01 [0.55–1.81] | 3,119 | 15.7 [11.4–21.2] | 18.2 [11.7–27.1] | 1.16 [0.67–1.93] |
| Gender<sup>4</sup> |  |  |  |  |  |  |  |  |
| Men | 63,264 | 3.8 [3.2–4.6] | 5.5 [4.6–6.5] | 1.42 [1.13–1.79] | 10,895 | 20.3 [17.4–23.6] | 32.6 [28.1–37.4] | 1.61 [1.34–2.01] |
| Women | 64,830 | 3.2 [2.7–3.8] | 4.8 [4.0–5.7] | 1.48 [1.18–1.91] | 9,733 | 18.6 [15.7–21.8] | 34.9 [29.9–40.2] | 1.88 [1.53–2.36] |
| Occupational social grade |  |  |  |  |  |  |  |  |
| ABC1 (more advantaged) | 80,353 | 3.1 [2.6–3.7] | 4.0 [3.4–4.6] | 1.28 [1.03–1.63] | 9,941 | 22.8 [19.5–26.4] | 34.5 [30.3–39.1] | 1.52 [1.28–1.91] |
| C2DE (less advantaged) | 48,235 | 4.0 [3.4–4.8] | 6.9 [5.7–8.2] | 1.70 [1.33–2.22] | 10,808 | 17.1 [14.5–20.1] | 34.1 [29.2–39.3] | 1.99 [1.59–2.46] |

<sup>1</sup> Unweighted sample size (number surveyed between July 2016 and April 2024).
<sup>2</sup> Data are weighted estimates of prevalence in the first and last months in the study period from logistic regression with survey month modelled non-linearly using restricted cubic splines. Trends across the entire period are shown in **Figure 1**.
<sup>3</sup> Prevalence ratio calculated as prevalence in April 2024 divided by prevalence in July 2016 with 95% CIs calculated using bootstrapping (1,000 replications).
<sup>4</sup> Sample sizes for men and women do not sum to the total because some participants identified in another way (adults $n=390$ ; smokers $n=100$ ) and some did not respond (adults $n=104$ ; smokers $n=21$ ).

Among adults who smoked, the proportion who also vaped increased from 19.5% to 34.2% (PR=1.76 [1.48–2.09]; **Table 1**). This increase was non-linear: dual use was relatively stable up to mid-2021, at an average of 18.6% [17.5–19.8%] of smokers between July 2016 and May 2021, then increased rapidly to 34.2% [30.9–37.8%] by April 2024 (**Figure 1A**).

**Figure 1.**
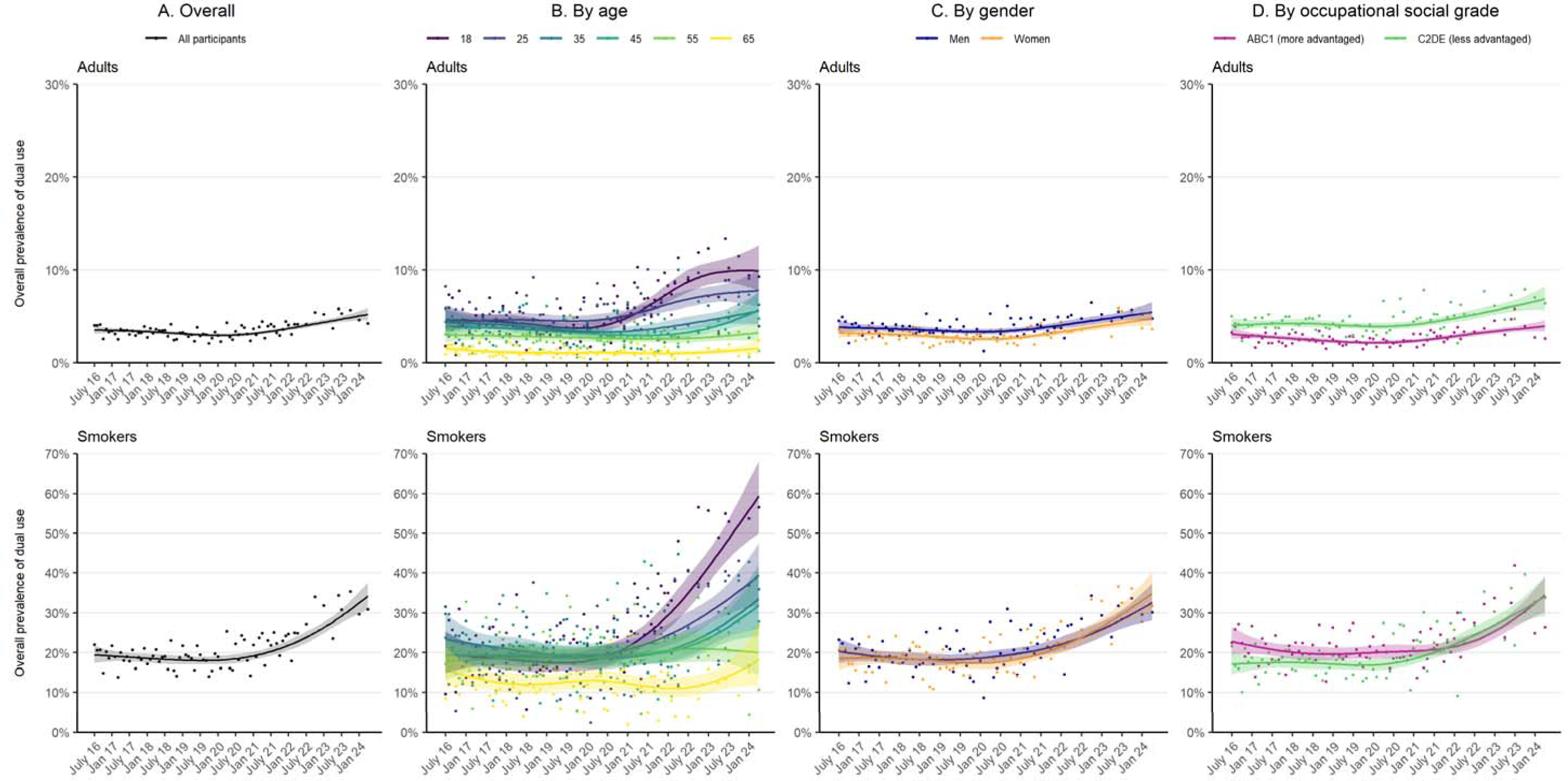
Trends in the overall prevalence of dual use of smoking and vaping in England, July 2016 to April 2024. Lines represent the modelled weighted proportion by monthly survey wave (modelled non-linearly using restricted cubic splines with five knots). Shaded bands represent 95% confidence intervals. Points represent the unmodelled weighted proportion by month. Sample sizes and modelled estimates of prevalence in the first and last months in the time series are reported in **Table 1**.

The increase in dual use was greatest among younger adults (increasing from 4.7% to 9.9% among 18-24-year-olds [19.6% to 59.4% among 18-24-year-old smokers]) and all but absent in the oldest group (1.5% to 1.5% among those aged ≥65 [15.7% to 18.2% among ≥65-year-old smokers]; **Figure 1B**). Time trends were similar by gender (**Figure 1C**) and occupational social grade (**Figure 1D**).

### Patterns of dual use, overall and by user characteristics

Of 4,247 dual users surveyed between July 2016 and April 2024, 3,744 (88.2%) provided data on how frequently they vaped (130 [3.1%] did not respond and 373 [8.8%] responded that they did not know). Among these participants, the most common pattern of dual use was daily cigarette smoking with daily vaping (49.0%) and the least common was non-cigarette tobacco smoking with non-daily vaping (1.5%; **Table 2**). Results were very similar when we excluded those who reported using other non-combustible nicotine products (*n*=599; **Table 2**).

**Table 2.**
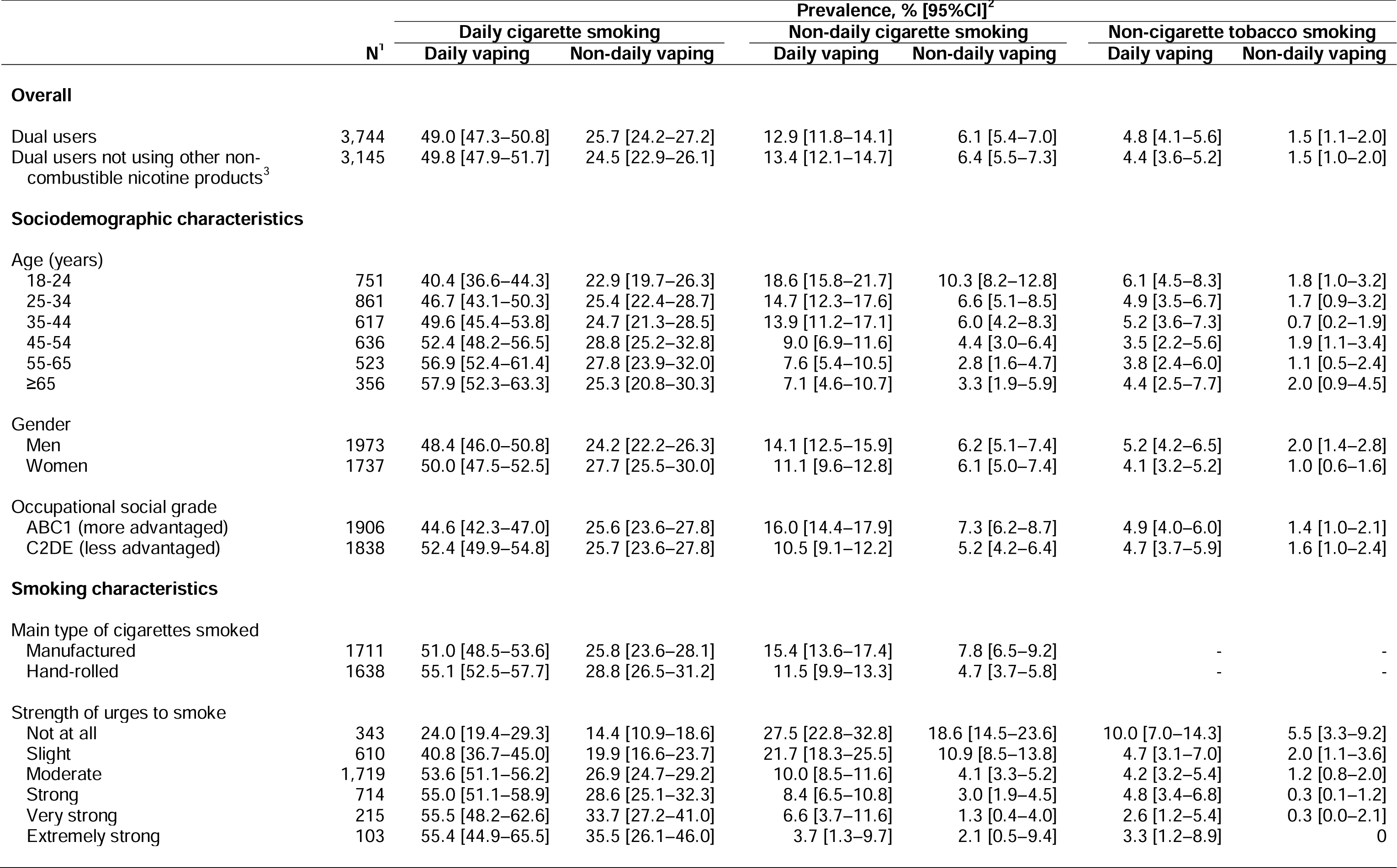
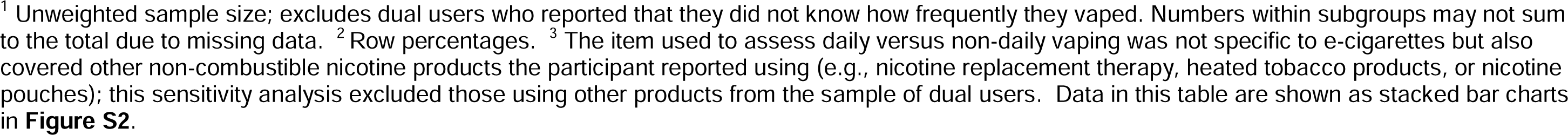
Patterns of dual use of smoking and vaping, overall and by sociodemographic and smoking characteristics (data aggregated across 2016-2024)

|  | N<sup>1</sup> | Prevalence, % [95%CI]<sup>2</sup> |  |  |  |  |  |
| --- | --- | --- | --- | --- | --- | --- | --- |
|  |  | Daily cigarette smoking |  | Non-daily cigarette smoking |  | Non-cigarette tobacco smoking |  |
|  |  | Daily vaping | Non-daily vaping | Daily vaping | Non-daily vaping | Daily vaping | Non-daily vaping |
| **Overall** |  |  |  |  |  |  |  |
| Dual users | 3,744 | 49.0 [47.3–50.8] | 25.7 [24.2–27.2] | 12.9 [11.8–14.1] | 6.1 [5.4–7.0] | 4.8 [4.1–5.6] | 1.5 [1.1–2.0] |
| Dual users not using other non-combustible nicotine products<sup>3</sup> | 3,145 | 49.8 [47.9–51.7] | 24.5 [22.9–26.1] | 13.4 [12.1–14.7] | 6.4 [5.5–7.3] | 4.4 [3.6–5.2] | 1.5 [1.0–2.0] |
| **Sociodemographic characteristics** |  |  |  |  |  |  |  |
| Age (years) |  |  |  |  |  |  |  |
| 18-24 | 751 | 40.4 [36.6–44.3] | 22.9 [19.7–26.3] | 18.6 [15.8–21.7] | 10.3 [8.2–12.8] | 6.1 [4.5–8.3] | 1.8 [1.0–3.2] |
| 25-34 | 861 | 46.7 [43.1–50.3] | 25.4 [22.4–28.7] | 14.7 [12.3–17.6] | 6.6 [5.1–8.5] | 4.9 [3.5–6.7] | 1.7 [0.9–3.2] |
| 35-44 | 617 | 49.6 [45.4–53.8] | 24.7 [21.3–28.5] | 13.9 [11.2–17.1] | 6.0 [4.2–8.3] | 5.2 [3.6–7.3] | 0.7 [0.2–1.9] |
| 45-54 | 636 | 52.4 [48.2–56.5] | 28.8 [25.2–32.8] | 9.0 [6.9–11.6] | 4.4 [3.0–6.4] | 3.5 [2.2–5.6] | 1.9 [1.1–3.4] |
| 55-65 | 523 | 56.9 [52.4–61.4] | 27.8 [23.9–32.0] | 7.6 [5.4–10.5] | 2.8 [1.6–4.7] | 3.8 [2.4–6.0] | 1.1 [0.5–2.4] |
| ≥65 | 356 | 57.9 [52.3–63.3] | 25.3 [20.8–30.3] | 7.1 [4.6–10.7] | 3.3 [1.9–5.9] | 4.4 [2.5–7.7] | 2.0 [0.9–4.5] |
| Gender |  |  |  |  |  |  |  |
| Men | 1973 | 48.4 [46.0–50.8] | 24.2 [22.2–26.3] | 14.1 [12.5–15.9] | 6.2 [5.1–7.4] | 5.2 [4.2–6.5] | 2.0 [1.4–2.8] |
| Women | 1737 | 50.0 [47.5–52.5] | 27.7 [25.5–30.0] | 11.1 [9.6–12.8] | 6.1 [5.0–7.4] | 4.1 [3.2–5.2] | 1.0 [0.6–1.6] |
| Occupational social grade |  |  |  |  |  |  |  |
| ABC1 (more advantaged) | 1906 | 44.6 [42.3–47.0] | 25.6 [23.6–27.8] | 16.0 [14.4–17.9] | 7.3 [6.2–8.7] | 4.9 [4.0–6.0] | 1.4 [1.0–2.1] |
| C2DE (less advantaged) | 1838 | 52.4 [49.9–54.8] | 25.7 [23.6–27.8] | 10.5 [9.1–12.2] | 5.2 [4.2–6.4] | 4.7 [3.7–5.9] | 1.6 [1.0–2.4] |
| **Smoking characteristics** |  |  |  |  |  |  |  |
| Main type of cigarettes smoked |  |  |  |  |  |  |  |
| Manufactured | 1711 | 51.0 [48.5–53.6] | 25.8 [23.6–28.1] | 15.4 [13.6–17.4] | 7.8 [6.5–9.2] | - | - |
| Hand-rolled | 1638 | 55.1 [52.5–57.7] | 28.8 [26.5–31.2] | 11.5 [9.9–13.3] | 4.7 [3.7–5.8] | - | - |
| Strength of urges to smoke |  |  |  |  |  |  |  |
| Not at all | 343 | 24.0 [19.4–29.3] | 14.4 [10.9–18.6] | 27.5 [22.8–32.8] | 18.6 [14.5–23.6] | 10.0 [7.0–14.3] | 5.5 [3.3–9.2] |
| Slight | 610 | 40.8 [36.7–45.0] | 19.9 [16.6–23.7] | 21.7 [18.3–25.5] | 10.9 [8.5–13.8] | 4.7 [3.1–7.0] | 2.0 [1.1–3.6] |
| Moderate | 1,719 | 53.6 [51.1–56.2] | 26.9 [24.7–29.2] | 10.0 [8.5–11.6] | 4.1 [3.3–5.2] | 4.2 [3.2–5.4] | 1.2 [0.8–2.0] |
| Strong | 714 | 55.0 [51.1–58.9] | 28.6 [25.1–32.3] | 8.4 [6.5–10.8] | 3.0 [1.9–4.5] | 4.8 [3.4–6.8] | 0.3 [0.1–1.2] |
| Very strong | 215 | 55.5 [48.2–62.6] | 33.7 [27.2–41.0] | 6.6 [3.7–11.6] | 1.3 [0.4–4.0] | 2.6 [1.2–5.4] | 0.3 [0.0–2.1] |
| Extremely strong | 103 | 55.4 [44.9–65.5] | 35.5 [26.1–46.0] | 3.7 [1.3–9.7] | 2.1 [0.5–9.4] | 3.3 [1.2–8.9] | 0 |

<sup>1</sup> Unweighted sample size; excludes dual users who reported that they did not know how frequently they vaped. Numbers within subgroups may not sum to the total due to missing data. <sup>2</sup> Row percentages. <sup>3</sup> The item used to assess daily versus non-daily vaping was not specific to e-cigarettes but also covered other non-combustible nicotine products the participant reported using (e.g., nicotine replacement therapy, heated tobacco products, or nicotine pouches); this sensitivity analysis excluded those using other products from the sample of dual users. Data in this table are shown as stacked bar charts in **Figure S2**.

Patterns of dual use were similar by gender but differed according to users’ age and occupational social grade (**Table 2**). Relative to older dual users, those who were younger were less likely to report daily cigarette smoking with daily vaping (e.g., 40.4% among 18-24s vs. 57.9% among ≥65s) and more likely to report non-daily cigarette smoking with daily/non-daily vaping (e.g., 28.9% vs. 10.4%; **Table 2**, **Figure S2A**). Relative to those who were less advantaged, dual users from more advantaged social grades were less likely to report daily cigarette smoking with daily vaping (44.6% vs. 52.4%) and more likely to report non-daily cigarette smoking with daily vaping (16.0% vs. 10.5%; **Table 2**, **Figure S2C**).

There were also differences by the main type of cigarettes smoked and strength of urges to smoke (**Table 2**). Relative to those who mainly smoked hand-rolled cigarettes, dual users who mainly smoked manufactured cigarettes were more likely to report non-daily cigarette smoking with daily/non-daily vaping (23.2% vs. 16.2%) and less likely to report daily cigarette smoking with daily/non-daily vaping (76.8% vs. 83.9%; **Table 2**, **Figure S2D**), although the latter comparisons were uncertain, as indicated by overlapping 95% CIs. Dual users who reported weaker urges to smoke were less likely to report daily cigarette smoking with daily/non-daily vaping (e.g., 38.4% among those reporting no urges in the past 24 hours vs. 90.9% among those reporting extremely strong urges) and more likely to report non-daily cigarette smoking with daily/non-daily vaping (e.g., 46.1% vs. 5.8%) or non-cigarette tobacco smoking with daily/non-daily vaping (e.g., 15.5% vs. 3.3%; **Table 2**, **Figure S2E**).

Patterns of dual use differed by vaping duration and purpose (**Table 3**). Dual users who had been vaping for longer were less likely to report daily cigarette smoking with daily vaping (e.g., 45.6% vs. 52.7% for those who had been vaping for >12 months vs. ≤6 months) and more likely to report non-daily cigarette smoking with daily vaping (e.g., 16.1% vs. 9.9%; **Table 3**, **Figure S2F**). Those who reported using e-cigarettes to cut down the amount they smoked or in situations when smoking is not permitted were more likely than those who reported vaping for other reasons to report daily/non-daily cigarette smoking with daily vaping (69.2% and 62.1% vs. 28.0%) and less likely to report daily/non-daily cigarette smoking with non-daily vaping (26.1% and 30.5% vs. 66.3%; **Table 3**, **Figure S2G**). There were no notable differences between those who had vs. had not unsuccessfully used e-cigarettes in a past-year quit attempt (**Table 3**, **Figure S2H**).

**Table 3.** Patterns of dual use of smoking and vaping, by vaping characteristics and harm perceptions of e-cigarettes vs. cigarettes (data aggregated across 2016-2024)

|  | N<sup>1</sup> | Prevalence, % [95%CI]<sup>2</sup> |  |  |  |  |  |
| --- | --- | --- | --- | --- | --- | --- | --- |
|  |  | Daily cigarette smoking |  | Non-daily cigarette smoking |  | Non-cigarette tobacco smoking |  |
|  |  | Daily vaping | Non-daily vaping | Daily vaping | Non-daily vaping | Daily vaping | Non-daily vaping |
| **Vaping characteristics** |  |  |  |  |  |  |  |
| Vaping duration |  |  |  |  |  |  |  |
| ≤6 months | 1,635 | 52.7 [50.1–55.4] | 27.0 [24.7–29.4] | 9.9 [8.4–11.6] | 6.8 [5.6–8.3] | 2.7 [2.0–3.6] | 0.9 [0.5–1.5] |
| >6 and ≤12 months | 572 | 47.8 [43.5–52.2] | 24.2 [20.7–28.2] | 13.7 [10.9–17.0] | 7.2 [5.2–9.9] | 4.8 [3.2–7.2] | 2.2 [1.2–4.0] |
| >12 months | 1,513 | 45.6 [42.9–48.3] | 24.4 [22.1–26.8] | 16.1 [14.2–18.2] | 4.8 [3.9–6.1] | 7.2 [5.8–8.8] | 1.9 [1.2–3.0] |
| Vaping purpose |  |  |  |  |  |  |  |
| Cutting down the amount smoked<sup>3</sup> | 2,438 | 55.8 [53.6–57.9] | 21.0 [19.3–22.8] | 13.4 [12.0–15.0] | 5.1 [4.2–6.1] | 3.7 [3.0–4.6] | 1.0 [0.7–1.6] |
| Use in situations when smoking is not permitted<sup>3</sup> | 2,548 | 48.9 [46.8–51.0] | 24.8 [23.0–26.7] | 13.2 [11.9–14.7] | 5.7 [4.8–6.8] | 5.5 [4.6–6.6] | 1.8 [1.3–2.5] |
| Other reasons | 40 | 25.9 [13.9–43.1] | 55.3 [38.6–71.0] | 2.1 [0.3–14.2] | 11.0 [3.7–28.5] | 4.5 [1.1–17.0] | 1.2 [0.2–8.5] |
| Used e-cigarettes in a past-year quit attempt |  |  |  |  |  |  |  |
| No | 2,552 | 48.4 [46.3–50.5] | 26.5 [24.7–28.4] | 12.0 [10.7–13.4] | 6.5 [5.6–7.6] | 4.9 [4.1–6.0] | 1.7 [1.2–2.3] |
| Yes | 1,192 | 50.3 [47.3–53.4] | 23.9 [21.4–26.7] | 14.8 [12.8–17.2] | 5.3 [4.1–6.8] | 4.4 [3.3–5.9] | 1.2 [0.7–2.1] |
| **Harm perceptions of e-cigarettes vs. cigarettes** |  |  |  |  |  |  |  |
| E-cigarettes are less harmful | 2,097 | 49.4 [47.1–51.7] | 22.3 [20.5–24.3] | 14.7 [13.2–16.5] | 6.1 [5.1–7.3] | 5.7 [4.7–7.0] | 1.7 [1.1–2.4] |
| E-cigarettes are equally harmful | 1,136 | 48.4 [45.2–51.5] | 28.9 [26.1–31.8] | 11.0 [9.2–13.2] | 6.7 [5.3–8.4] | 3.7 [2.7–5.1] | 1.3 [0.7–2.3] |
| E-cigarettes are more harmful | 228 | 41.0 [34.4–48.1] | 37.0 [30.5–44.1] | 8.4 [5.4–12.9] | 7.3 [4.3–12.0] | 3.4 [1.6–6.9] | 2.8 [1.2–6.7] |
| Don't know | 283 | 55.4 [49.1–61.6] | 27.2 [22.1–33.1] | 10.9 [7.4–15.8] | 3.1 [1.6–6.0] | 3.0 [1.3–6.7] | 0.3 [0.0–2.1] |

<sup>1</sup> Unweighted sample size; excludes dual users who reported that they did not know how frequently they vaped. Numbers within subgroups may not sum to the total due to missing data. <sup>2</sup> Row percentages. <sup>3</sup> These purposes were not mutually exclusive ( $n=1,438$ reported vaping both to cut down the amount smoked and to use in situations when smoking is not permitted). Data in this table are shown as stacked bar charts in **Figure S2**.

Patterns of dual use also differed by participants’ perceptions of the relative harms of e-cigarettes vs. cigarettes (**Table 3**). Across groups of smokers (i.e., daily cigarette, non-daily cigarette, non-cigarette tobacco), dual users who thought e-cigarettes were less than or equally as harmful as cigarettes, or who were unsure, were more likely to vape daily than non-daily whereas those who thought e-cigarettes were more harmful were similarly likely to vape daily or non-daily (**Table 3**, **Figure S2I**). Dual users who believed e-cigarettes were less harmful were the most likely to report non-daily cigarette smoking and daily vaping (14.7%) and those who thought e-cigarettes were more harmful were most likely to report daily cigarette smoking and non-daily vaping (37.0%). Those who were unsure of the relative harms of the two products were the most likely to report daily cigarette smoking with daily vaping (55.4%).

### Trends in patterns of dual use

Patterns of dual use changed over the study period (**Figure 2**). Between July 2016 and April 2024, the proportion of dual users reporting daily cigarette smoking with non-daily vaping fell by more than 50% (from 35.2% to 15.0%; PR=0.43 [0.29–0.63]), offset primarily by an increase in the proportion reporting non-daily cigarette smoking with daily vaping (from 7.6% to 21.5%; PR=2.84 [1.71–4.72]; **Table 4**, **Figure 2B**).

**Figure 2.**
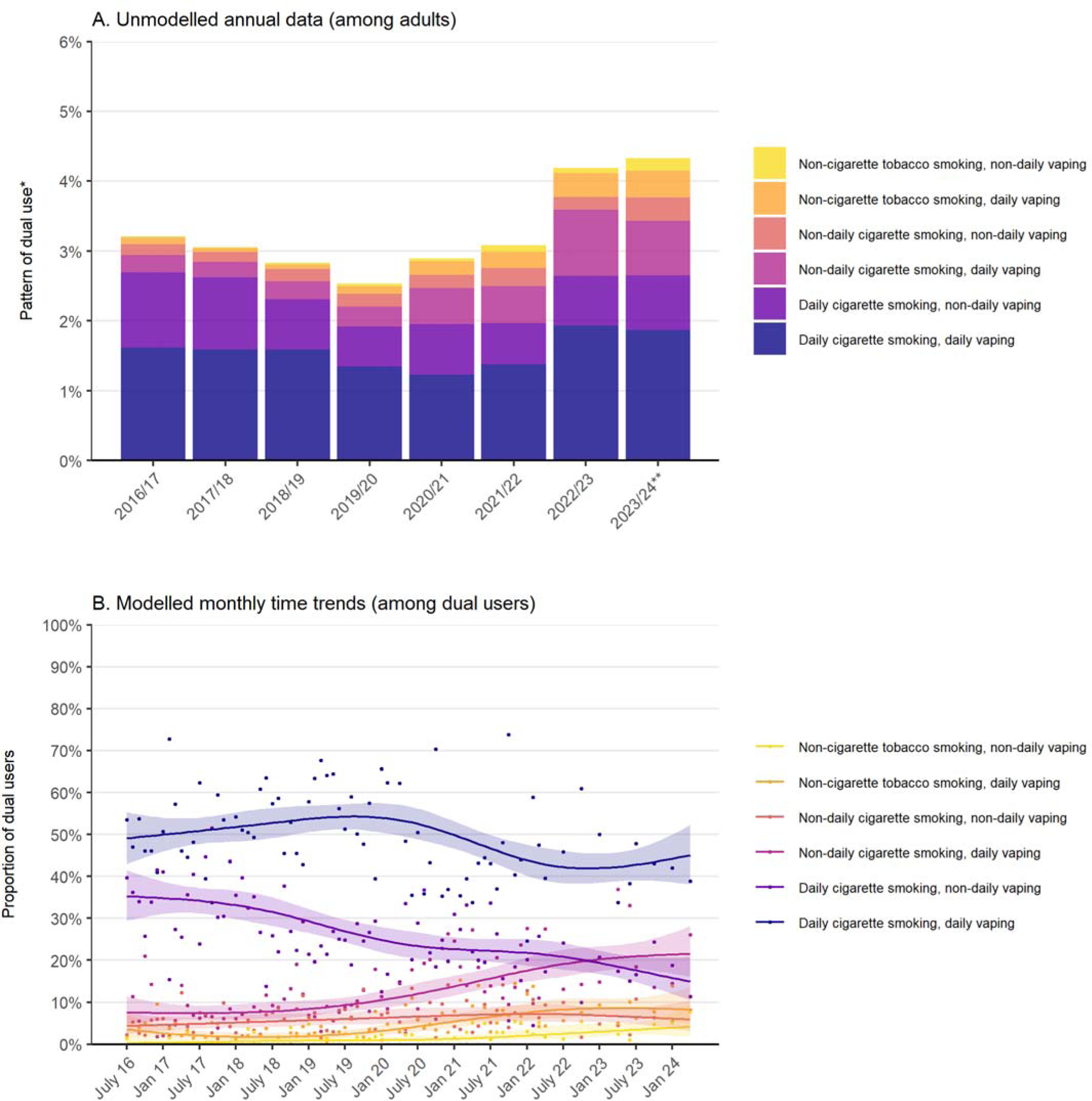
Trends in different patterns of dual use of smoking and vaping in England, 2016 to 2024. Panel A shows the proportion of adults who reported each pattern of dual use, aggregated by survey year (July-June). *Sample excludes dual users who reported that they did not know how frequently they vaped; **Figure S3** shows data including these participants. **2023/24 only includes data up to April. Corresponding figures for smokers and dual users are provided in **Figure S4**. Panel B shows modelled time trends in the proportion of dual users who reported each pattern of dual use. Lines represent the modelled weighted proportion by monthly survey wave (modelled non-linearly using restricted cubic splines with five knots). Shaded bands represent 95% confidence intervals. Points represent the unmodelled weighted proportion by month. Modelled estimates of prevalence in the first and last months in the time series are reported in **Table 4**.

**Table 4.**
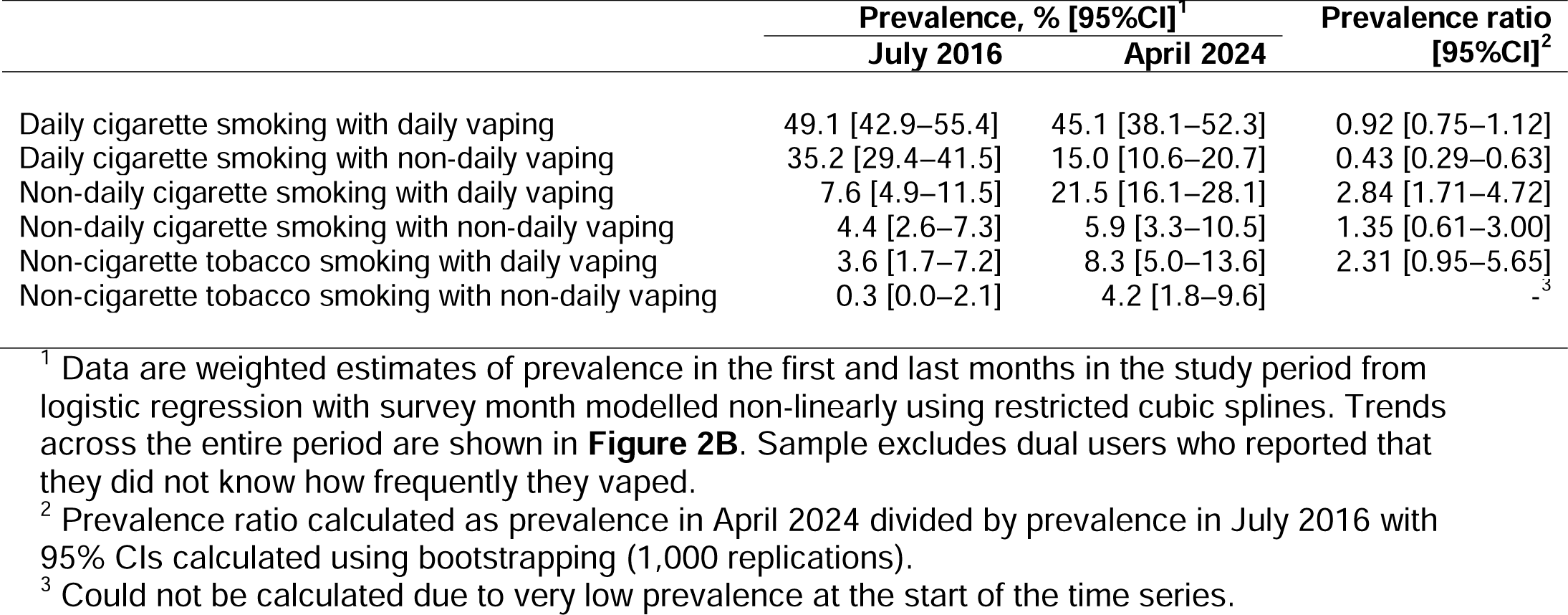
Modelled estimates of patterns of smoking and vaping among dual users (*n*=4,247) in July 2016 and April 2024.

There were also large but uncertain increases in the proportions reporting non-cigarette tobacco smoking with daily vaping (from 3.6% [1.7–7.2%] to 8.3% [5.0–13.6%]) and non-cigarette tobacco smoking with non-daily vaping (from 0.3% [0.0–2.1%] to 4.2% [1.8–9.6%]; **Table 4**, **Figure 2B**). Collapsed across daily and non-daily vaping, the increase in the proportion of dual users reporting non-cigarette tobacco smoking with vaping increased three-fold (from 3.7% [1.8–7.2%] to 12.4% [8.1–18.6%]; PR=3.38 [1.52–7.49]).

The most prevalent form of dual use – daily cigarette smoking with daily vaping – remained relatively unchanged as a proportion of dual users (from 49.1% in July 2016 to 45.1% in April 2024; **Table 4**). However, because the overall prevalence of dual use increased (**Figure 1A**), there was an absolute increase in the proportion of adults who reported daily use of both cigarettes and e-cigarettes in recent years (from 1.2% [1.1–1.3%] in 2020/21 to 1.9% [1.5–2.2%] in 2023/24; **Figure 2A**).

As of April 2024, 45.1% of dual users reported daily cigarette smoking with daily vaping, 21.5% non-daily cigarette smoking with daily vaping, 15.0% daily cigarette smoking with non-daily vaping, 8.3% non-cigarette tobacco smoking with daily vaping, 5.9% non-daily cigarette smoking with non-daily vaping, and 4.2% non-cigarette tobacco smoking with non-daily vaping (**Table 4**).

## Discussion

Across the study period, there was an increase in the proportion of smokers in England who vaped, particularly among younger age groups. This increase was non-linear, with little change between 2016 and mid-2021, followed by a rapid increase up to April 2024. This mirrors recent trends in adult vaping prevalence, which has risen substantially since new disposable e-cigarettes started to become popular.^11,19^ The inverse age gradient in the increase in dual use is also consistent with recent increases in vaping being greater at younger ages and virtually absent among those aged ≥65.^11^ As of April 2024, one in 20 adults in England, and one in three smokers, reported dual use of e-cigarettes and combustible tobacco; these numbers were much higher among 18-24-year-olds (one in 10 and more than one in two, respectively).

Across the entire period, the most common pattern of dual use was daily cigarette smoking with daily vaping. This differs from previous evidence from the 2016 ITC Survey, which indicated the most common pattern in England was daily smoking with non-daily vaping.^12^ This discrepancy may be due to changes in the way people have opted to dual use over time, or changes in the types of people who are dual using. We found the proportion of dual users reporting daily cigarette smoking with non-daily vaping decreased considerably between 2016 and 2024. This was offset primarily by an increase in the proportion reporting non-daily cigarette smoking with daily vaping, alongside smaller increases in the proportions smoking non-cigarette tobacco with vaping daily or non-daily. These changes in patterns of dual use may be explained by the population of dual users comprising a greater proportion of younger adults over time, who were more likely than older dual users to report non-daily cigarette smoking with daily vaping or exclusively smoking non-cigarette tobacco.

The combination of daily cigarette smoking and daily vaping was more common among dual users who were older, less advantaged, mainly smoked hand-rolled cigarettes, and had stronger urges to smoke. This is not surprising, given these variables are typically associated with greater dependence on smoking.^17,20^ Daily cigarette smoking with daily vaping was also more common among those who reported vaping to cut down or in situations where smoking was not permitted (vs. other reasons) and those who had been vaping for shorter periods (e.g., ≤6 months vs. >12 months), while non-daily cigarette smoking with daily vaping was more common among those who had been vaping for more than a year. These findings are consistent with the possibility that vaping, even outside of a formal quit attempt, may support people to transition away from smoking, possibly by initially reducing their daily cigarette consumption then moving to non-daily smoking.^21^

Dual users who thought e-cigarettes were less or equally harmful to cigarettes, or were unsure, were more likely to vape daily than non-daily, whereas those who thought e-cigarettes were more harmful were similarly likely to vape daily or non-daily. Evidence suggests that while daily vaping is associated with increased attempts to quit smoking and increased success in quitting, people who vape non-daily are not more likely to quit than those who do not vape at all.^22^ Therefore, misperceptions of e-cigarettes as more harmful than cigarettes could potentially deter dual users from vaping in a way that could help them to move towards quitting smoking.

Our findings have several implications for research, policy, and practice. First, there is a need for more research on longitudinal transitions to assess the extent to which new cohorts of people in England who are dual using follow the path from daily to non-daily smoking and non-daily to daily vaping to eventual cessation of combustibles. Second, targeted interventions may be warranted to encourage more older smokers to try vaping. Third, better education is needed about the likely health impacts of different patterns of dual use; in particular, the importance of moving away from daily smoking (given this has remained stable when dual using with daily vaping). Finally, accurate harm perceptions of e-cigarettes vs. cigarettes may be important for encouraging dual users to move from daily to non-daily smoking.

This study had several limitations. The data collected were cross-sectional and participants were not asked which product they started using first, so we could not determine temporality of uptake of smoking and vaping. It is likely that older participants will have started with cigarettes, but this is less certain for younger age groups. The assessment of smoking status distinguished between those who smoked cigarettes and those who exclusively used other forms of combustible tobacco but did not identify whether cigarette smokers also smoked non-cigarette tobacco. In addition, the frequency of smoking was not assessed among those who reported exclusively smoking non-cigarette tobacco, so we were unable to distinguish between daily and non-daily use. The item used to determine daily vs. non-daily vaping not only assessed how frequently participants used e-cigarettes but also any other non-combustible nicotine products they reported using; however, a sensitivity analysis that excluded those using multiple products produced a very similar pattern of results. Finally, there were also some missing data on vaping frequency (largely due to participants responding that they did not know how frequently they vaped) which meant we could not determine the pattern of dual use for a subset of participants.

In conclusion, the overall prevalence of dual use increased non-linearly to approximately 5% of adults in England in 2024. The proportion of smokers who also vape increased rapidly since 2021, which was when disposable e-cigarettes started to become popular. There has been a shift in patterns of dual use away from more frequent smoking towards more frequent vaping. This may be the result of increasing prevalence of dual use among younger adults, who are more likely than older dual users to smoke non-daily and vape daily. Dual users who had been vaping for longer were more likely to smoke non-daily and vape daily. Those who perceived the harms of vaping as being greater than the harms of smoking were less likely to vape daily, indicating misperceptions around the relative harms of vaping and smoking could undermine the potential benefits of dual use for smoking cessation.

## Data Availability

All data produced in the present study are available upon reasonable request to the authors

## Declarations

### Ethics approval

Ethical approval for the STS was granted originally by the UCL Ethics Committee (ID 0498/001). The data are not collected by UCL and are anonymised when received by UCL.

