## Supplementary material for "Trends and patterns of dual use of combustible tobacco and e-cigarettes among adults in England: a population study, 2016-2024": Table S1

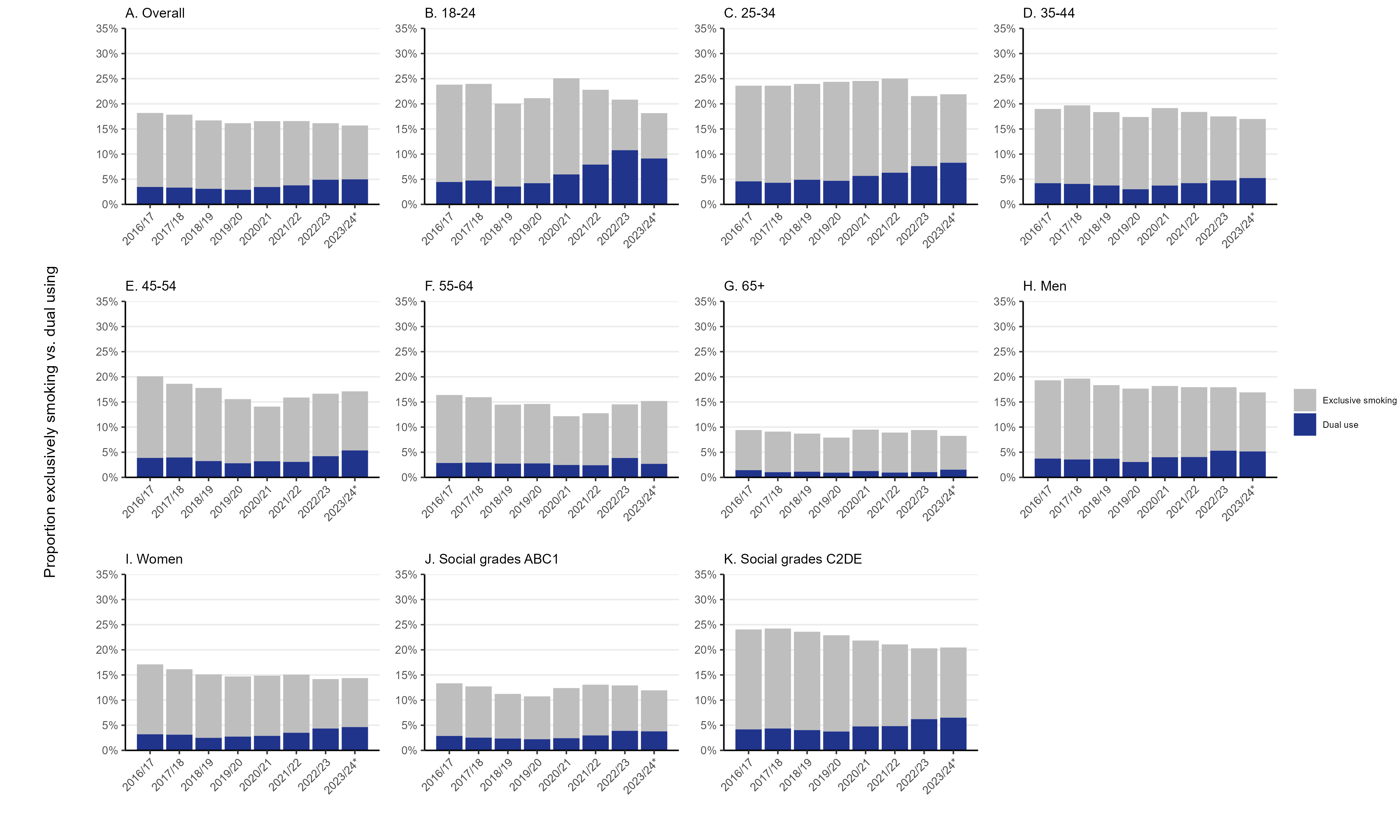

### Figure S1. Prevalence of dual use of smoking and vaping vs. exclusive smoking in England, 2016 to 2024, overall and by sociodemographic characteristics. Proportion of adults in England who report dual use vs. exclusive smoking, aggregated by survey year (July-June), (A) overall and by age (B-G), gender (H-I), and occupational social grade (J-K). ABC1 = more advantaged, C2DE = less advantaged. *2023/24 only includes data up to April.

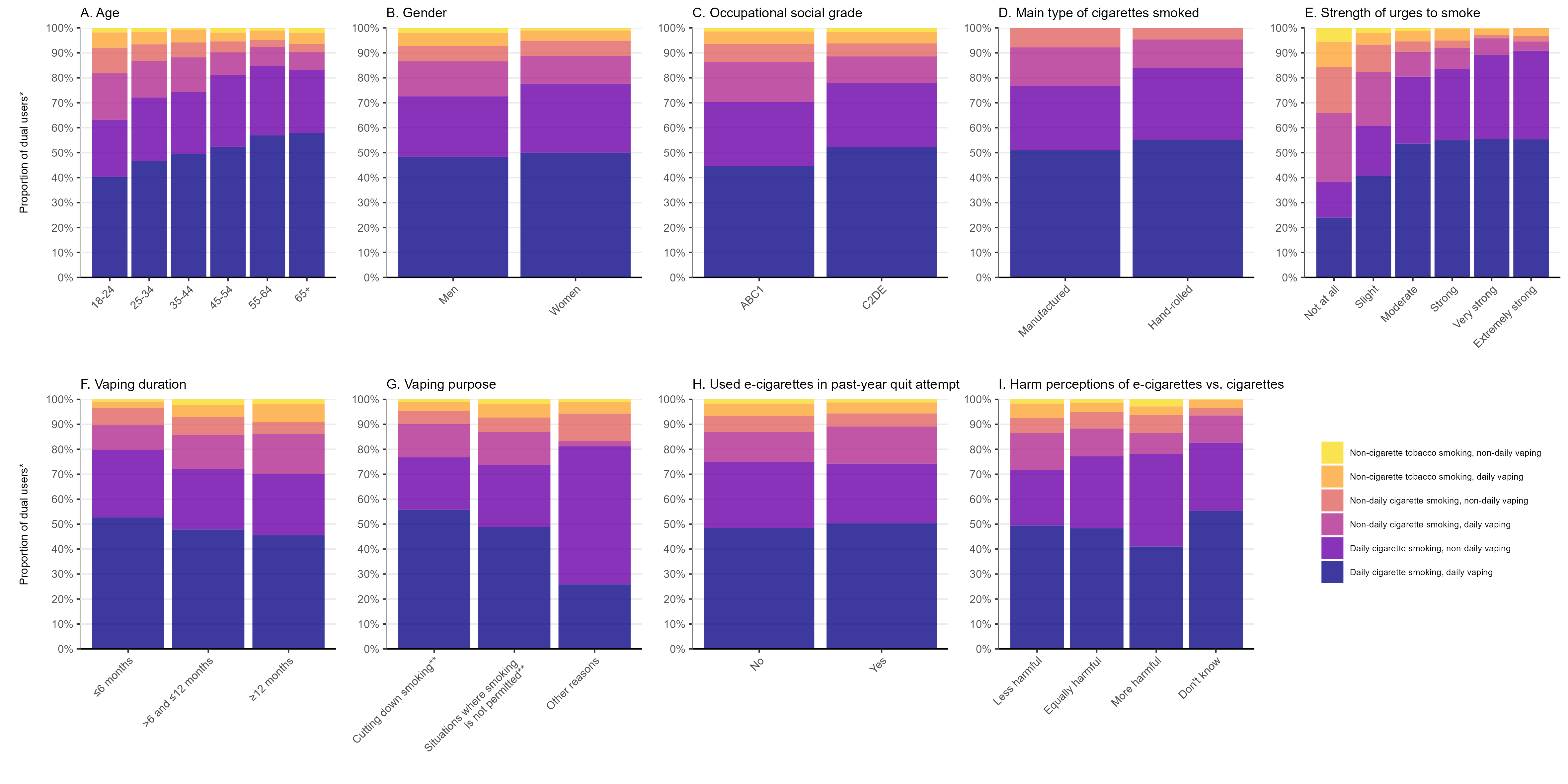

### Figure S2. Patterns of dual use of smoking and vaping in England, by user characteristics. Proportion of dual users who reported each pattern of dual use, by sociodemographic, smoking, and vaping characteristics and harm perceptions of e-cigarettes vs. cigarettes. *Sample excludes dual users who reported that they did not know how frequently they vaped. **These purposes were not mutually exclusive.

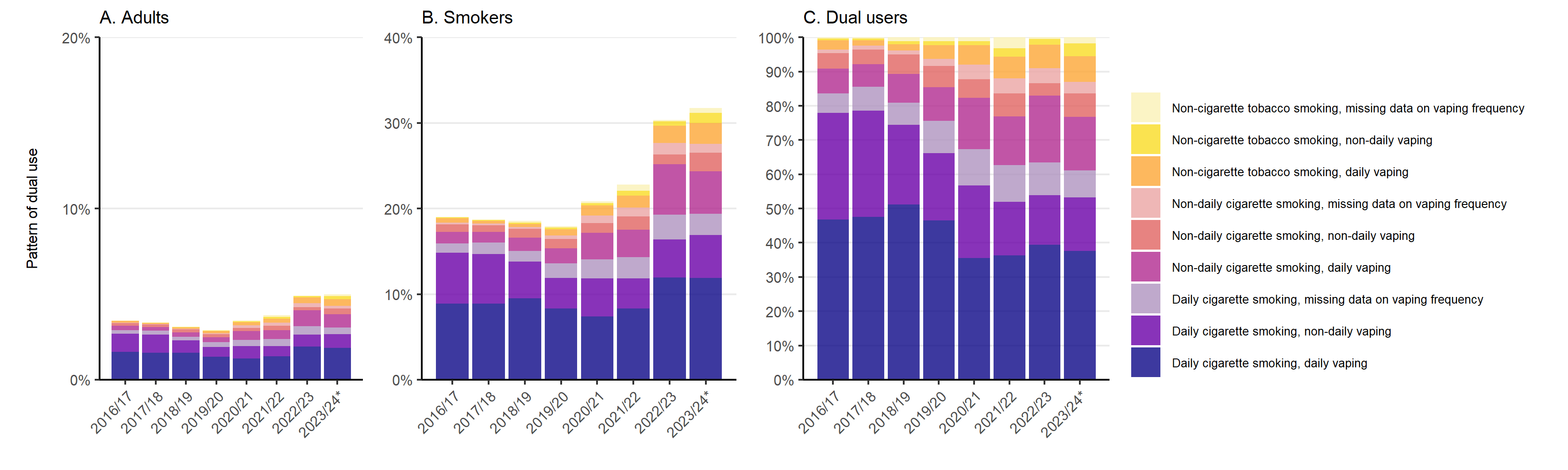

### Figure S3. Trends in different patterns of dual use of smoking and vaping in England, 2016 to 2024 – including dual users with missing data on vaping frequency. Proportion of (A) adults, (B) smokers, and (C) dual users who reported each pattern of dual use, aggregated by survey year (July-June), including dual users who reported that they did not know how frequently they vaped. **2023/24 only includes data up to April.

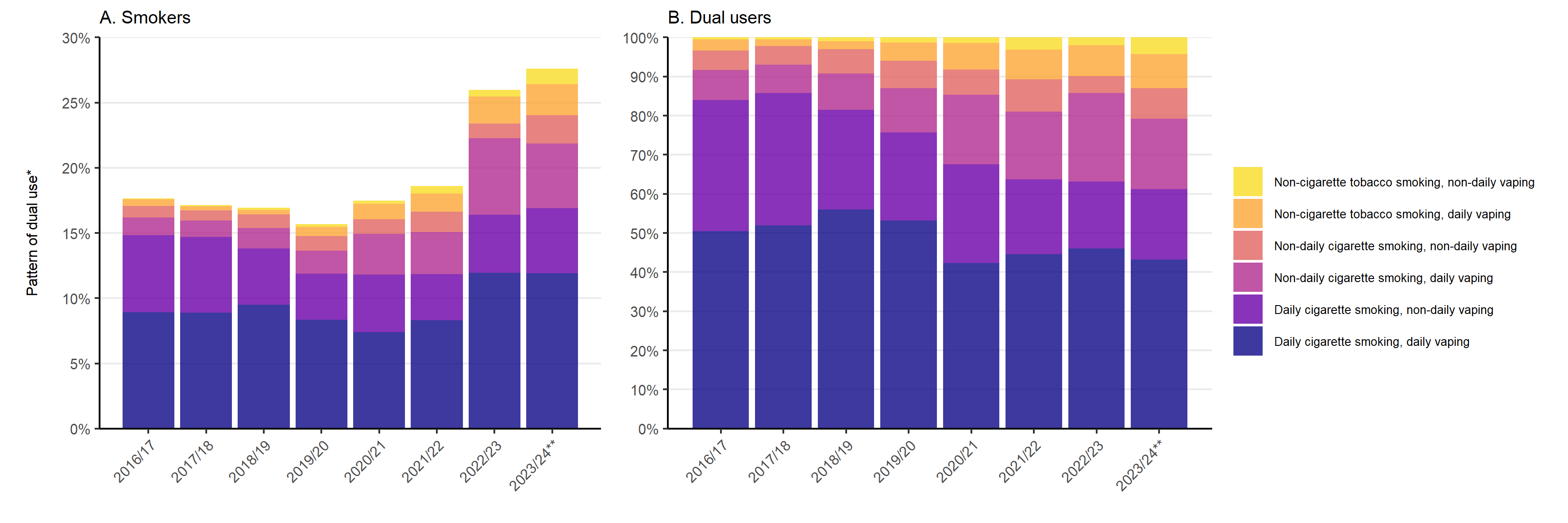

### Figure S4. Trends in different patterns of dual use of smoking and vaping among smokers and dual users in England, 2016 to 2024. Proportion of (A) smokers and (B) dual users who reported each pattern of dual use, aggregated by survey year (July-June). * Data are not shown for dual users who reported that they did not know how frequently they vaped; Figure S3 shows data including these participants. **2023/24 only includes data up to April.
